# The course of the UK COVID 19 pandemic; no measurable impact of new variants

**DOI:** 10.1101/2021.03.16.21253534

**Authors:** D Ellis, D Papadopoulos, S Mukherjee, U Ukwu, N Chari, K Charitopoulos, I Donkov, S Bishara

## Abstract

**Introduction:** In November 2020, a new SARS-COV-2 variant or the ‘Kent variant’ emerged in the UK, and became the dominant UK SARS-COV-2 variant, demonstrating faster transmission than the original variant, which rapidly died out. However, it is unknown if this altered the overall course of the pandemic as genomic analysis was not common place at the outset and other factors such as the climate could alter the viral transmission rate over time. We aimed to test the hypothesis that the overall observed viral transmission was not altered by the emergence of the new variant, by testing a model generated earlier in the pandemic based on lockdown stringency, temperature and humidity.

**Methods:** From 1/1/20 to 4/2/21, the daily incidence of SARS-COV-2 deaths and the overall stringency of National Lockdown policy on each day was extracted from the Oxford University Government response tracker. The daily average temperature and humidity for London was extracted from Wunderground.com.

The viral reproductive rate was calculated on a daily basis from the daily mortality data for each day. The correlation between log_10_ of viral reproductive rate and lockdown stringency and weather parameters were compared by Pearson correlation to determine the time lag associated with the greatest correlation.

A multivariate model for the log10 of viral reproductive rate was constructed using lockdown stringency, temperature and humidity for the period 1/1/20 to 30/9/20. This model was extrapolated forward from 1/10/20 to 4/2/21 and the predicted viral reproductive rate, daily mortality and cumulative mortality were compared with official data.

**Results:** On multivariate linear regression, the optimal model had and R^2^ 0f 0.833 for prediction of log_10_ viral reproductive rate 13 days later in the model construction period, with (coefficient, probability) lockdown stringency (−0.0109, p=0.0000), humidity (0.0038, p=0.0041) and temperature (−0.0035, p=0.0008). When extrapolated to the validation period (1/10/20 to 4/2/21), the model was highly correlated with daily (Pearson coefficient 0.88, p=0.0000) and cumulated SARS-COV-2 mortality (Pearson coefficient 0.99, p=0.0000).

**Conclusion:** The course of the SARS-COV-2 pandemic in the UK seems highly predicted by an earlier model based on the lockdown stringency, humidity and temperature and unaltered by the emergence of a newer viral genotype.

## Introduction

Since the Sars-CoV-2 outbreak was declared a global pandemic by the World Health Organisation in March 2020, there have been over 109 million confirmed cases and 2.4 million deaths worldwide.[1] In the United Kingdom, there have been more than 118,000 deaths and 4 million cases,[2], [3] though the true number of cases is likely to be much higher due to asymptomatic infections. Seroprevalence studies prior to widespread vaccination suggest up to 1 in 8 would have tested positive for Sars-CoV-2 antibodies.[4]

In the absence of highly effective treatments or a vaccine, the initial response to the pandemic centred on preventing person to person transmission through social distancing or lockdown methods. The implementation of lockdowns slowed the spread of the pandemic in many nations, though as seen in the last great pandemic, the Spanish Flu in 1918[5], multiple waves of infection have occurred.

The international Lockdown response has been meticulously documented by the Oxford University Government Response tracker[6] which consists of individual lockdown and economic policies scored on an ordinal scale and a composite lockdown score which measures the overall lockdown strength, on a daily basis, for almost every country in the world; a sizeable undertaking which has improved our understanding of the effectiveness of lockdown strategies.

A number of studies have demonstrated that Sars-CoV-2 transmission is higher in cold weather.[7] Humidity may be an important factor, with a negative correlation to Sars-CoV-2 transmission and deaths shown in a number of studies.[8], [9] We have applied the methodology described in a previous study[10], which demonstrated the means of predicting the viral reproductive rate based on lockdown and weather parameters.

Rapid genomic sequencing has never previously been available on the scale seen in the Covid-19 pandemic and provides insight into the disease pathology and a means of assessing existing and potential therapies. The SARS-COV-2 was first sequenced in January 2020. The viral RNA sequence is a single positive RNA of 29811 nucleotides in length[11] and significantly different from MERs-CoV or SARs-CoV. The evolution rate is thought to be typical of an RNA virus at 10^−4^ substitutes per base per year.[12]

The UK, through the efforts of the COG-UK consortium, has sequenced more Covid-19 genomes than any other country.[13] The B.1.1.7, or 20I/501Y.V1 or ‘UK’ or ‘Kent’ variant[14] was first detected on the 20/09/20 and was sequenced early in October 2020.[15] It spread in low levels throughout the population in November but has now become the predominant UK variant.[16] It was declared a particular variant of concern by Public Health England on the 14th of December 2020.[16], [17] The number of spike protein mutations raised the possibility that vaccine susceptibility would be reduced,[18] though data suggests that the Oxford/AstraZeneca vaccine appears to about 74 per cent effective at preventing symptomatic infections due to B.1.1.7, compared with 85 per cent for other variants.[19]: A further E484K mutation may confer greater resistance to existing antibodies and vaccines, though only a handful of cases have been identified in the UK.[20] B.1.1.7 has been associated with a higher Case fatality rate by a factor of 1.35 compared to the wild type according to data from Scientific Advisory group for Emergencies in UK[21] and 70% increased transmissibility versus the original strain.[16]

The aim of this study is to determine the effect of the emergence of the Kent variant on the overall progression of the pandemic in the UK, through the application of a predictive model based on lockdown, and temperature and humidity in keeping with the methodology of a previous study[10]

## Methods

National record of lockdown as determined by the composite stringency index was obtained from the Oxford SARS-COV-2 Government Response Tracker for the UK, from 1/1/20 to 4/02/21. The composite is an average of 9 government responses scored on individual ordinal scales and ranges from 0 for no Lockdown to 100 for the most stringent Lockdown. This was converted to a 7-day rolling mean lockdown.

Daily mean temperature and humidity records were extracted from the website wunderground.org for London (deemed to be representative of the whole UK) for the study period (1/1/20 to 04/02/21).

The official UK daily mortality data was also extracted from Oxford SARS-COV-2 Government Response Tracker. The Viral reproductive (R_0_) rate was calculated for each date by dividing the number of people who died due to Sars-CoV-2 in the following 6 days by the number of people who had died in the preceding 6 days, in keeping with infective period of 6 days and an incubation period of 6 days, similar to the method described by Heald.[22] This observed R_0_ was corrected for the effect of immunity by dividing the observed R_0_ by the proportion of the population that were susceptible. The effect of population immunity was factored by taking the infection fatality rate of Sars-CoV-2 infection as 0.5%,[6] and this was used to calculate the prevalence of prior infections in the community. Immunity was assumed to be homogenous and lasting across the population, and prevented both death and transmission. The portion of the population that was susceptible was recalculated each day as ((5,000-deaths per million))/5,000)). A 7-day rolling average corrected R_0_ was calculated and converted to a log base 10.

The relationship between temperature, humidity and lockdown was assessed by the Pearson correlation by comparing 7-day rolling average weather parameter and average lockdown with the corrected log10 viral reproductive rate for 0 to 28 days after that date. The timing of the peak association between weather parameters and the effect on corrected Log10 Viral reproductive rate (LR) was assessed, by maximisation of the Pearson’s coefficient.

The study period was divided into a test period (The first 274 days from 1/1/20 to 30/09/2020) and assessment period (From 01/10/20 to 04/02/21, 127 days). Multivariate linear regression was applied to data from the study test period, combining 7 day rolling stringency index, 7 day rolling mean temperature and 7-day mean humidity for predictive parameters for LR in the following 0 to 35 days, and the model with the highest R^2^ was selected for the assessment period.

The multivariate equation from the test period (with the highest R^2^) was applied to the assessment period using the same coefficients and time lag in the determination of LR. The calculated LR was converted to an observed LR by multiplying the R_0_ by the proportion susceptible using the method above and this was used to calculate the predicted rolling active infections and then predicted daily deaths and cumulative deaths. The correlation between the actual and predicted observed LR, daily deaths and cumulative deaths was determined by Pearson coefficient. The regression equation was used to calculate the predicted lockdown stringency required to keep R_0_ below 1 for the whole period (1/1/20 to 4/2/21).

All statistics were calculated using NCSS 2021. Statistical significance was taken as p<0.05 throughout.

## Results

Figure 1: Univariate correlation between log_10_ UK viral reproductive rate (uncorrected for immunity) and a) London 7-day mean rolling temperature and 7-day mean rolling humidity and b) 7-day mean rolling lockdown stringency, according to differing time lags after parameter measurement (n=209). Higher humidity is associated with an increased R_0_ 0-28 days later, with a peak at 20 days, Pearson coefficient 0.5963 (p=0.000). Temperature had a negative correlation for most of the test period but had a positive association after 21 days. 7 day rolling lockdown had a strong negative correlation, peaking at 12 days later, Pearson coefficient −0.8856 (p=<0.0001).

**Figure 1.**
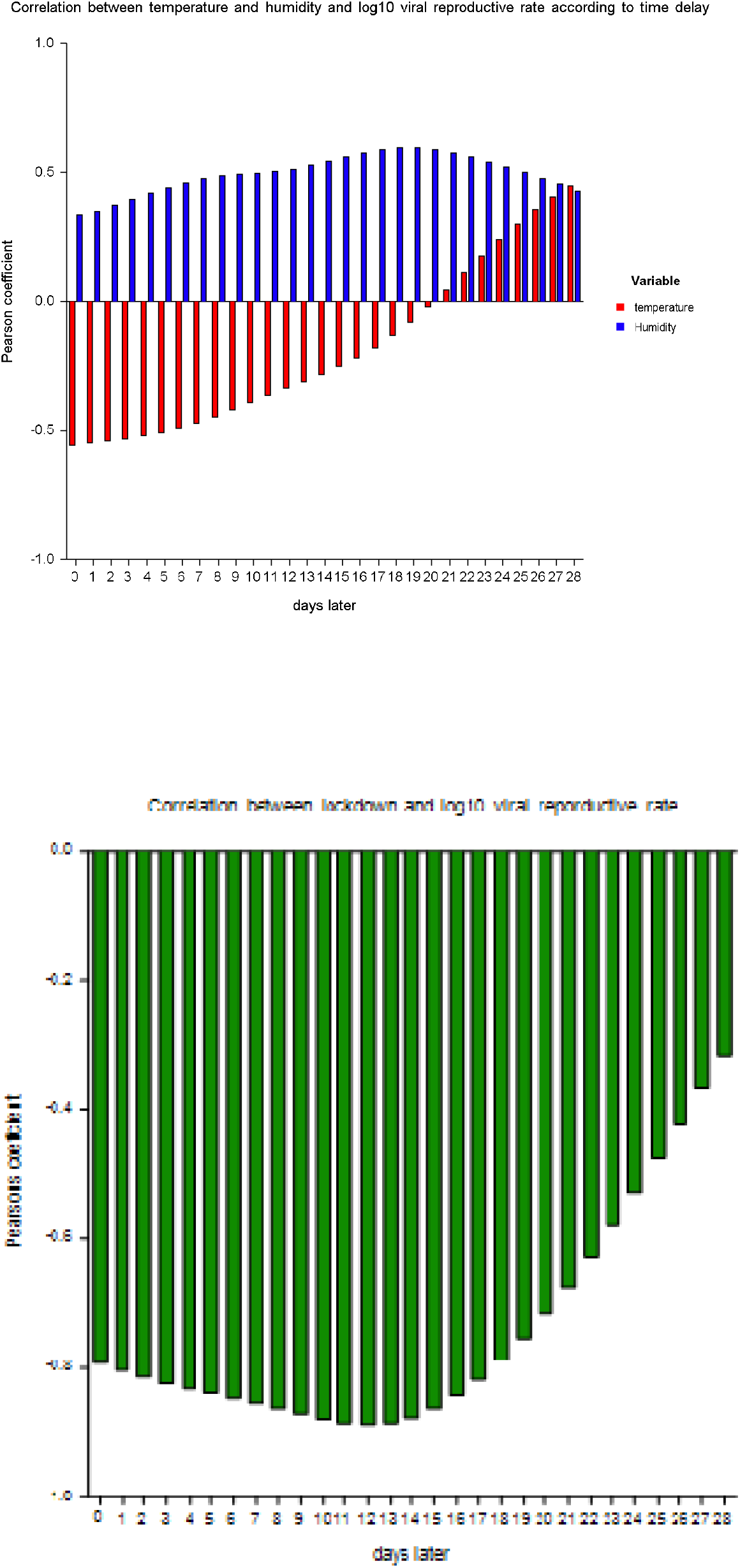
a) Correlation between 7 day rolling temperature and humidity and log10 viral reproductive rate according to differing time lags (n=209); b) Correlation between 7 day rolling lockdown stringency and log10 viral reproductive rate according to differing time lags (n=209).

Table 1, shows the results of multivariate regression, assessing the combination of a 7-day rolling lockdown, 7-day rolling humidity and 7-day rolling temperature in predicting log_10_viral reproductive rate (uncorrected for immunity). The strongest combination model was for 13 days later, R^2^=0.833, (p<0.0001, n=209). Lockdown is the strongest independent predictor with a strong negative correlation. Humidity was significantly positively correlated and temperature was significantly negatively correlated.

**Table 1;.** multiple regression model used for prediction in model of UK pandemic. The model predicts log_10_Viral reproductive rate (uncorrected for immunity), 13 days after the parameter measurement. R^2^=0.833

| Independent Variable | Regression Coefficient | Standard Error | Standardized Coefficient | T-Statistic to Test | Probability Level |
| --- | --- | --- | --- | --- | --- |
|  | **b(i)** | **Sb(i)** | **Coefficient** | **H0: $\beta(i)=0$** | **Level** |
| **Intercept** | 0.7586 | 0.1172 | 0 | 6.471 | 0 |
| **Temperature** | -0.0035 | 0.001 | -0.1069 | -3.407 | 0.0008 |
| **Humidity** | 0.0038 | 0.0013 | 0.099 | 2.902 | 0.0041 |
| **Lockdown** | -0.0109 | 0.0005 | -0.8051 | -22.31 | 0 |

Figure 2. The model generated from 1/1/20 to 30/09/20 was extrapolated forward to 4/2/21. Shown is a comparison of the observed and predicted viral reproductive rate, daily mortality and cumulative mortality illustrated as a time series and correlation graph (now all corrected for immunity). In the time series, the predicted viral reproductive rate tracks the actual rate. The measured correlation seems low with Pearson coefficient of 0.20 (95% CI 0.02-0.36, p=0.020), as the magnitude of the variability of R_0_ in the evaluation period is large relative to the size of the R_0_, but 65% of the daily estimates of the predicted viral rate are within 0.2 of the actual observed rates. The variables derived from the calculated R_0_ are more precise than the R_0,_ showing a Pearson coefficient of 0.88 for prediction of daily mortality (95% CI 0.83 −0.91, p<0.0001) and cumulative mortality 0.99 (95% CI 0.98-0.99). The predicted cumulative mortality for 4/2/21 was 105554 which is close to official cumulative mortality for that date of 110250.

**Figure 2;.**
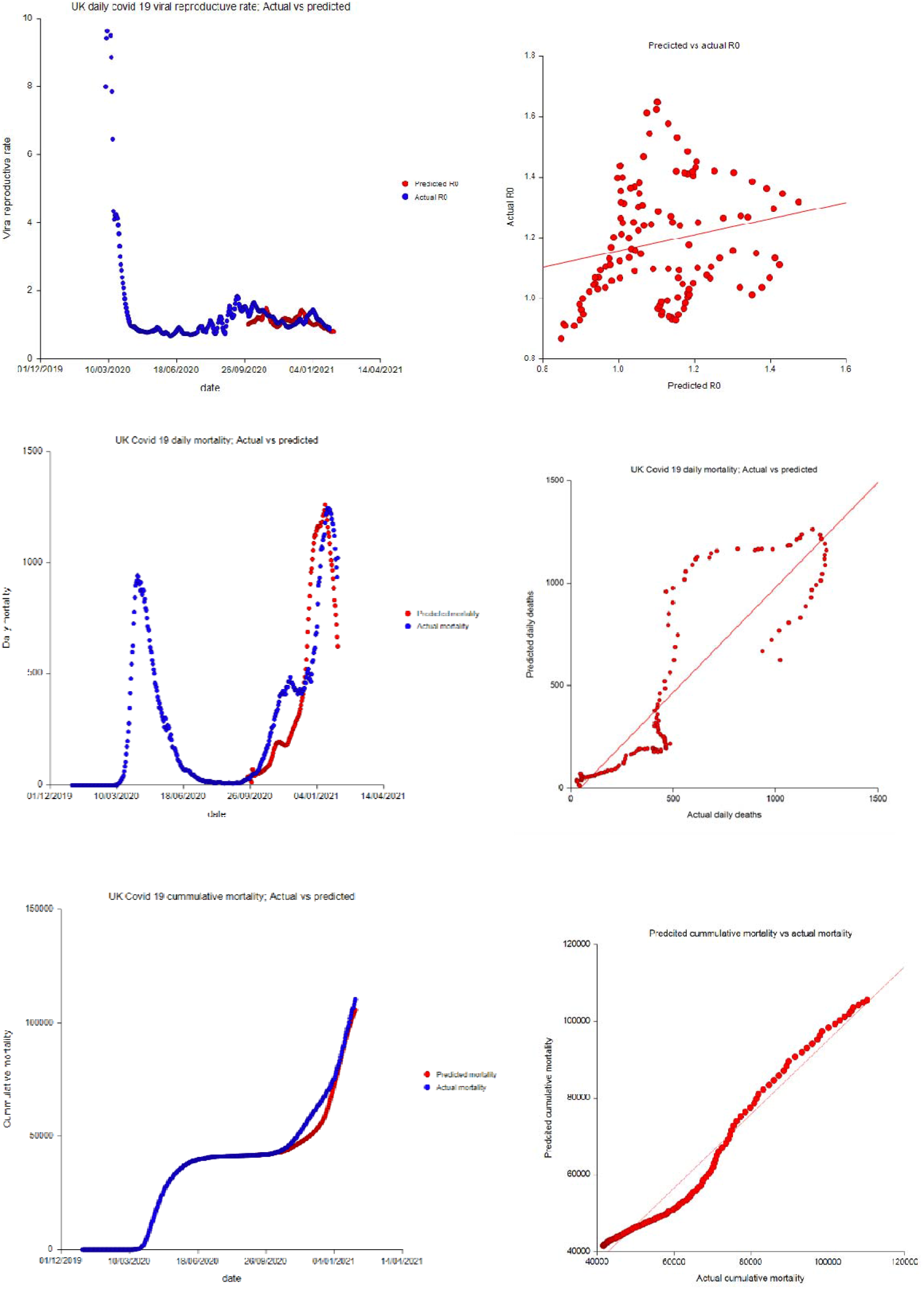
Comparison between actual and predicted (corrected for immunity) R_0_, daily deaths and cumulative deaths shown as a time series and scatter graph with regression line. Correlation coefficients are 0.20 for R_0_ (95% CI 0.02-0.36, p=0.020) 0.88, (95% CI 0.83-0.91, p=0.000) for daily deaths and 0.99, (95% CI 0.98-0.99, p=<0.001) for cumulative deaths (n=121).

Figure 3 demonstrates the effect of the weather parameters in the prediction model on the required strength of lockdown to bring the observed viral reproductive rate (factoring immunity) to 1. This predicted lockdown stringency is compared to the actual lockdown stringency. The effect of weather is more modest than the lockdown but the predicted lockdown stringency varies from about 60 in the summer to 80 for the winter months. For the entire period after April 2020 the UK lockdown stringency has been > 60 but there are periods where the actual stringency has tracked above or below the required stringency.

**Figure 3.**
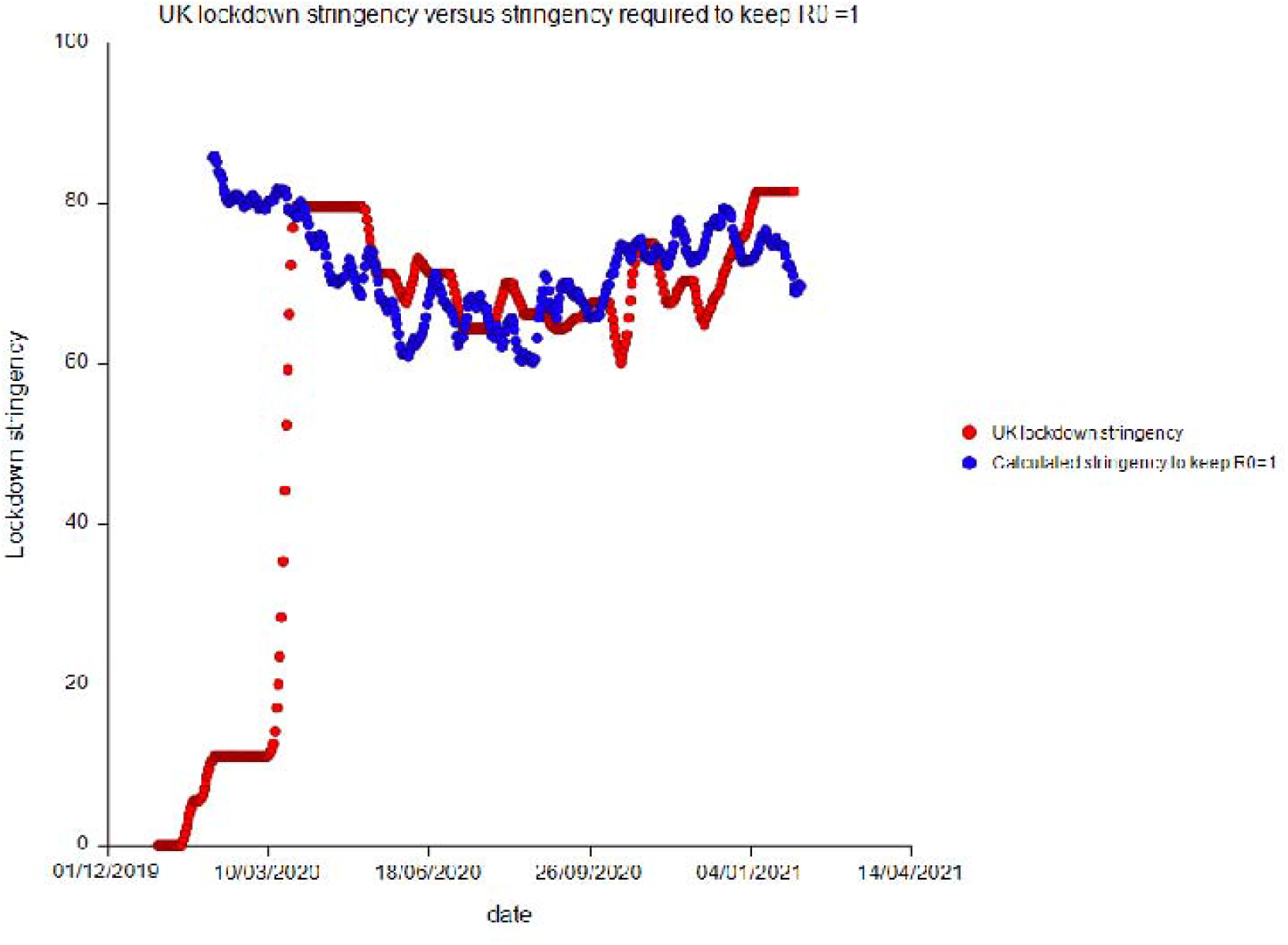
Comparison between UK actual lockdown stringency and that calculated as required for a(corrected for immunity) R_0_=1.

## Discussion

This model assesses the effect of lockdown and weather parameters on the Sars-CoV-2-19 viral reproductive rate, demonstrating the size of their effects and the time delay which determines these relationships. Of the three parameters evaluated, the lockdown was most predictive of the viral reproductive rate, with a peak of association at 12 days. This is in keeping with the published mean incubation period of 5.8 days, and median period from symptom onset to death of 7 days.[23], [24]

Increased humidity was associated with increased viral transmission with peak univariate association at 20 days, perhaps reflecting fomite transmission which could be relatively delayed. It is important to note that positive correlation has not been universally reported, including in a previous application of this model to multiple countries.[10] Some studies show a bell-shaped curve conferring lower virus survival at moderate levels of humidity,[25] potentially explained by the behaviour of salts dissolved within the respiratory droplets.[26]

Temperature had a negative association with transmission. This aligns with other studies of transmission and is to be expected due to the widely documented inactivation of coronaviridae at higher temperatures[27], [28], and reduced lifespan of respiratory droplets.[26]

The model generated here is similar to a previous proposed model,[10] the previous model could not be extrapolated due to a small change in the generation of the lockdown stringency score. The newer model is generated over a longer period of time (9 months versus 4 months) but also extrapolated over a longer period (4 months versus 1.5 months). It seems to perform well in that the predicted cumulative mortality for 4/2/21 was 105554 which is close to official cumulative mortality for that date of 110250. The model, which was extrapolated from the 1/10/2020 predicted a peak daily mortality of 1262 on the 16/1/2021 which is very close to the actual peak 7 day rolling (centred on the day) mortality of 1248 on the 20/1/2021. The effect of vaccination programme, which began on the 8^th^ of December 2020, will have minimal impact on the data here as only 2.28 million people (3.4%) were vaccinated by 10^th^ of January and it takes about 3 weeks for vaccines to have an effect on transmission and longer on mortality.[29]

The estimation of immunity is an important feature of the model which we have estimated from the cumulative mortality as being associated with an IFR of 0.5%. Thus the final population immunity on 4/2/21 was estimated to be 31.6% which is higher than 25% of people[30] who are estimated to have antibodies in the 28 days prior to 11/2/21, though the effect of T-cell immunity remains an unknown factor. The factoring of immunity has a huge impact on the model output, particularly on the later viral reproductive rates. If the effect of immunity from prior infection is removed, then the model predicts that there would have 700000 deaths, and 140 million infections equivalent to everyone in the UK having Covid 19 twice.

The pandemic seems to have progressed numerically in a manner that is highly predicted by a prior model that is based only on temperature, humidity lockdown stringency and projected immunity. The predicted viral rate seems to be unaffected by the emergence of the B.1.17 variant, as in the model increased R_0_ is predicted by the decreased temperature and relatively reduced lockdown stringency. How this fit with observations that the new variant has flourished whilst the original variant has receded, and side by side comparisons have demonstrated increased transmissibility and CFR. A possible explanation for the comparative increased CFR is the finding, that in a prolonged infection, the virus can evolve to the new strain within an individual,[31] and it may be that the patients who were sicker and more likely to die, had poorer immunes systems and higher viral loads, were more likely to develop the new strain even if they weren’t originally infected with the new strain, when its overall prevalence in the UK was low.

Whilst genomics is a powerful tool in assessing, its use in the population was not as widely practiced in the early phase of the pandemic and the latter-day sub classification and renaming and un-shaming of mutational variations overlooks that the virus has been constantly evolving from day 1 and there is some balance between the heterogeneous and incompletely understood immune response and the virus’s ability to adapt. In the study immunity was treated in a simplistic all or nothing manner but in reality, there is likely a spectrum of immunity between newer and older variants which may provide a survival advantage. It has also been demonstrated that mutations that helped avoid the immune system may lead to a defect in infectivity that is counterbalanced by the emergence of further mutations.[31] New variants may only become more infective due a relatively increased immunity to an older variant.

Another possible explanation is that the response tracker only document official government mandates. Even in the absence of governmental instructions, in disease resurgence or publicity surrounding a more dangerous variant, a proportion of individuals are likely to implement their own measures such as avoiding busy locations and enhanced hygiene practices.[10] This element of ‘personal lockdown’ may also account for the discrepancy between transmissibility of the Kent variant and the results here, though overall our data demonstrates a very strong correlation with the lockdown stringency score.

Figure 3 demonstrates the actual UK lockdown versus the level required to supress the viral reproductive rate to 1. For both the first and second peak there is a relative delay initiating lockdown tightening followed by a prolonged tighter lockdown. The R_0_ during the extrapolation period ranged from 0.86 to 1.65. To reverse the effect of an R_0_ of 1.65 for 1 week would require an R_0_ of 0.61 for 1 week which was unachievable through UK lockdown, with the levels of immunity seen in the extrapolation period, in fact it would require 3 weeks of lockdown at 0.86 to reverse the effects of 1 week of R_0_ at 1.65 on the daily infection rate. It is easier to prevent the peak from developing by early modest tightening than flattening it by delayed more stringent measures. Some have proposed short intermittent lockdowns, as means of maximising the time out of stringent lockdown without allowing the pandemic to grow during each lockdown cycle.[32] As Covid 19 seems to be becoming endemic, lockdown only serve to delay the inevitable spread throughout the population unless there is an alternative means of providing immunity and fortunately this seems to be achievable by the current vaccination programme.

Weaknesses of this study include use of temperature from the capital city which may vary widely across regions and the limitations of recorded Sars-CoV-2 data. While the use of deaths as the primary endpoint is likely to be the most accurate measure for a large population, it does increase the time between observed parameters and outcomes. The infection fatality rate is also assumed to be static, but is likely to decrease in response to improved treatment and novel therapies, such as the use of Dexamethasone.[20]

Calculations using the viral reproductive rate are compound in nature, and a small variation in the number of infections at one point in time will have a large bearing on the number of infections at a later date. However, we believe any predictive model should centre on prediction of the viral reproductive rate, which though widely variable, is predicted by the variables evaluated here, in particular the degree of lockdown. As the focus of investigation turns more to emergent variants, this model could benefit the understanding of their effects in the context of the pandemic as a whole.

## In conclusion

The overall course of the Covid-19 pandemic in the United Kingdom does not seem to have been significantly altered by the emergence of the Kent variant of the disease, despite its selective advantage over previous dominant strains.

## Data Availability

Data is available upon request

